# Coping mechanisms of students with mental disorders at Busitema University, an exploratory qualitative study at Mbale and Busia campuses

**DOI:** 10.1101/2023.06.12.23290962

**Authors:** Enid Kawala Kagoya, Joseph Mpagi, Paul Waako, Martha R L Muhwezi, Agnes Namaganda, Allan.G. Nsubuga, Christine Etoko Atala, Francis Okello, Ambrose Okibure, Ronald Kibuuka, Ernest Wandera, Kalisiti Ndamanywa, Joseph Kirabira

**Affiliations:** Institute of Public Health, Department of Community Health, Busitema University, Faculty of Health Sciences, P.O Box 1460 Mbale, Uganda; Deans Office, Department of Academics, Research and Innovation, Busitema University, Faculty of Health Sciences, P.O Box 1460 Mbale, Uganda; Department of Psychiatry, Busitema University, Faculty of Health Sciences, P.O Box 1460 Mbale, Uganda; Department of Physiology, Makerere University, College of Health Sciences, P.O Box 7062, Kampala, Uganda; Executive Director, Forum for Africa Women Educationalists (FAWE) Africa, P.O Box 24117, Kampala, Uganda; Office vice chancellor, Busitema University, P.O Box 1460, Busia Uganda; Department of Anesthesia, Mbarara University of Science and Technology, P.O. Box 1410, Mbarara Uganda

**Keywords:** Mental disorder, coping strategies, university, students, Uganda

## Abstract

**Background:** Over the past years, Ugandan universities have been grappling with mental health issues among students. The prevalence of mental disorders was up to 60% among undergraduate students in Uganda. This trend has been observed at many universities as mental health issues pose significant academic and social problems for many students. This study aimed to explore the coping mechanisms used by students with mental disorders at Busitema University.

**Methods:** Forty-two key informant interviews were conducted among students diagnosed with mental disorders based on Mini International Neuropsychiatric Interview. All the interviews followed a well-structured interview guide which was developed by the research team. All participants consented during the study. All interviews were audiotaped and recordings were later subjected to verbatim transcription. Each transcript was read several times by the research team. Thematic analysis was done following a deductive approach. Nvivo software was used to support the coding and grouping of codes.

**Results:** The study revealed several key coping strategies embraced by students experiencing mental health issues at the two campuses. Five themes emerged during the data analysis which included Alcohol and substance use, Use of sedatives, social and personal initiatives, seeking counselling services, withdrawal and confrontation and faith healing. A lot of subthemes emerged as indicated in table (2). Among the many were resorting to drug, alcohol and substance use, withdrawal, confrontations, witchcraft, engaging in co-curricular activities and other social events, abortions, seeking counselling and other forms of psychosocial support.

**Conclusions:** **In the face of increasing** mental health challenges among university students, their coping strategies amidst such situations need to be looked through and given much attention. Some are self-thought-through strategies while others are university-led interventions. There is a need to reinforce the good coping mechanisms and counter the bad ones for the betterment of the mental health status of these students.

## Introduction

The World Health Organization (WHO) defines health as the complete physical, mental and social well-being of people and not merely the absence of disease or any form of infirmity, in this case, the mental well-being of people is a priority as it affects the productivity of the population(1). Mental health is an important element in overall health and well-being, which encompasses intellectual health, spiritual health and emotional health(3). The World Psychiatric Association (WPA) proposes four standards of mental health: physical and mental coordination, adaptation to the social environment, well-being, and bringing one’s ability into full play in work(4). In Uganda today, most of the students in institutions of higher learning are struggling with mental health challenges yet there are no systematic coping strategies in place, instead, everyone comes up with their own depending on what the problem is and how it manifests(2). According to the WHO, the mental health of university students and other students in institutions of higher learning requires stable mood, coordinated interpersonal relations, objective self-judgment, and psychological adaption(1). Students at universities face more challenges on their own when they transition from adolescence to adulthood while leaving home to attend universities(5). The transition is often associated with an increase in depression, anxiety and stress however their coping strategies of mechanisms are one matter of concern(2).Therefore, understanding the coping mechanisms or strategies embraced by students is necessary to design interventions that can help to reduce the high prevalence of mental health problems plaguing students(6).

According to a survey conducted by the WHO, it indicated that over 30% of college students from eight countries were estimated to have experienced mental distress, which seriously affected their study and daily lives, leading to role impairments and poor academic records(4). In most universities, students adopt an array of mechanisms to deal with mental health issues varying from religious support(7), positive reinterpretation(8), active coping(9), planning, and use of instrumental support(10). Other strategies include breathing exercises(11), regular visits to a counsellor, talking to someone, temporary distraction, social networking, frequent exercising, healthy eating, meditation, joining a club, mindfulness, to calming strategies(12). While some students show resilience and resourcefulness in adapting effective coping strategies, approaches such as disengagement, isolation, over-indulgence, grieving and internalized coping strategies often pose negative consequences to the adolescent’s physical health while also aggravating existing mental health conditions(13)

Mental health is on the periphery of health policy and developmental agenda as strategies to promote enabling and nurturing environments for optimal mental health are still lacking(13). This is evident by the scarcity of epidemiological and large-scale data on the magnitude of undergraduate mental health problems, and coping strategies(14). Stigmatization of mental illness, coupled with a lack of innovative psychosocial interventions and counselling therapy, as well as limited human and technical capacity to deliver mental health services is contributing to the hesitancy of students in seeking help as well as their negative attitude toward counselling services(9). Not very many studies have been done to understand the coping strategies of undergraduate students with mental health challenges. It’s on this basis therefore that we decided to conduct a study to explore and understand the various coping strategies used by undergraduate students faced with mental health challenges at the two campuses of Busitema University that is Tororo and Mbale compasses to investigate the coping strategies adopted and the support systems offered to help deal with the problems.

## Materials and Methods

### Study Designs

This was a descriptive qualitative study among students at Busitema University faculties of Health Sciences and Engineering.

### Study Site

The sites of consideration were Busitema University Faculty of Health Sciences (Mbale Campus) and Busitema University Faculty of Engineering (Busitema main campus). Busitema University Faculty of Health Sciences is housed under Mbale Regional Referral Hospital in the Mbale district of Eastern Uganda which serves patients from 16 districts. It mainly offers medical courses at the undergraduate and postgraduate levels which include Bachelor’s in Medicine and Surgery, nursing, and Anesthesia. The second setting was Busitema University, Faculty of Engineering which is located in Busitema, Tororo district, and mainly offers Engineering courses at diploma and bachelor’s levels such as mechanical and civil engineering among others.

### Participants

The study included undergraduate students enrolled in Busitema University, either at the faculty of health sciences or the faculty of engineering who were diagnosed with one or more mental disorders using the Mini International Neuropsychiatric Interview. We excluded any student who had any severe or emergency medical condition that made them unable to respond meaningfully to the interview questions.

### Data Collection, instruments and Analysis

A total of 42 key informant interviews are conducted among students diagnosed with mental disorders. Students were approached during their free time through their leaders. Purposive sampling was used whereby students were first screened for mental disorders including (depression, anxiety, bipolar and psychotic disorder) using the Mini International Neuropsychiatric Interview. Participants who scored positive for any of the disorders were then considered for qualitative interview and selection was based on severity and number of mental disorders. The selected potential participants were contacted and those who consented were given an appointment for the qualitative interview. All the interviews followed a semi-structured interview guide which was developed by the research team. All participants consented during the study. All interviews were audiotaped and recordings were later subjected to verbatim transcription. Each transcript was read several times by the research team. Thematic analysis was done following a deductive approach. Nvivo software was used to support the coding and grouping of codes.

### Ethical Consideration

Ethical approval was sought and obtained from the Research Ethics Committees (REC) of Busitema University, the faculty of Health Sciences BUFHS-2022-11 and the National Council for Science and Technology (UNSCT-Number: HS2700ES). All interviews were conducted in safe and secure places with only one student at a time to ensure privacy and confidentiality of shared information. Administrative clearance was sought from the vice chancellor’s office to collect data from the students at both campuses. All methods and procedures were performed in accordance to all relevant national and international guidelines for conducting research involving human participants during COVID-19 pandemic.

## Results

A total of 42 students were interviewed. Among those interviewed were 9 males and 11 females from the faculty of health sciences as well as 15 males and 7 females were from the faculty of engineering (see Table 1)

**Table 1.** Social demographics of participants.

| Faculty | Male | Female | 1<sup>st</sup><br>year | 2<sup>nd</sup> year | 3<sup>rd</sup> year | 4<sup>th</sup> Year | 5<sup>th</sup> year |  |
| --- | --- | --- | --- | --- | --- | --- | --- | --- |
| **Health Sciences** | 9 | 11 | 5 | 5 | 6 | 1 | 3 | 20 |
| **Engineering** | 15 | 7 | 6 | 4 | 6 | 6 |  | 22 |
| **Total** | 24 | 18 | 11 | 9 | 12 | 7 | 3 | 42 |

Participants reported coping mechanisms that fall mainly in five thematic areas which include; alcohol and other substance use, social and personal initiatives, seeking counselling services, withdrawal or disengagement and faith healing (see Table 2).

**Table 2.**
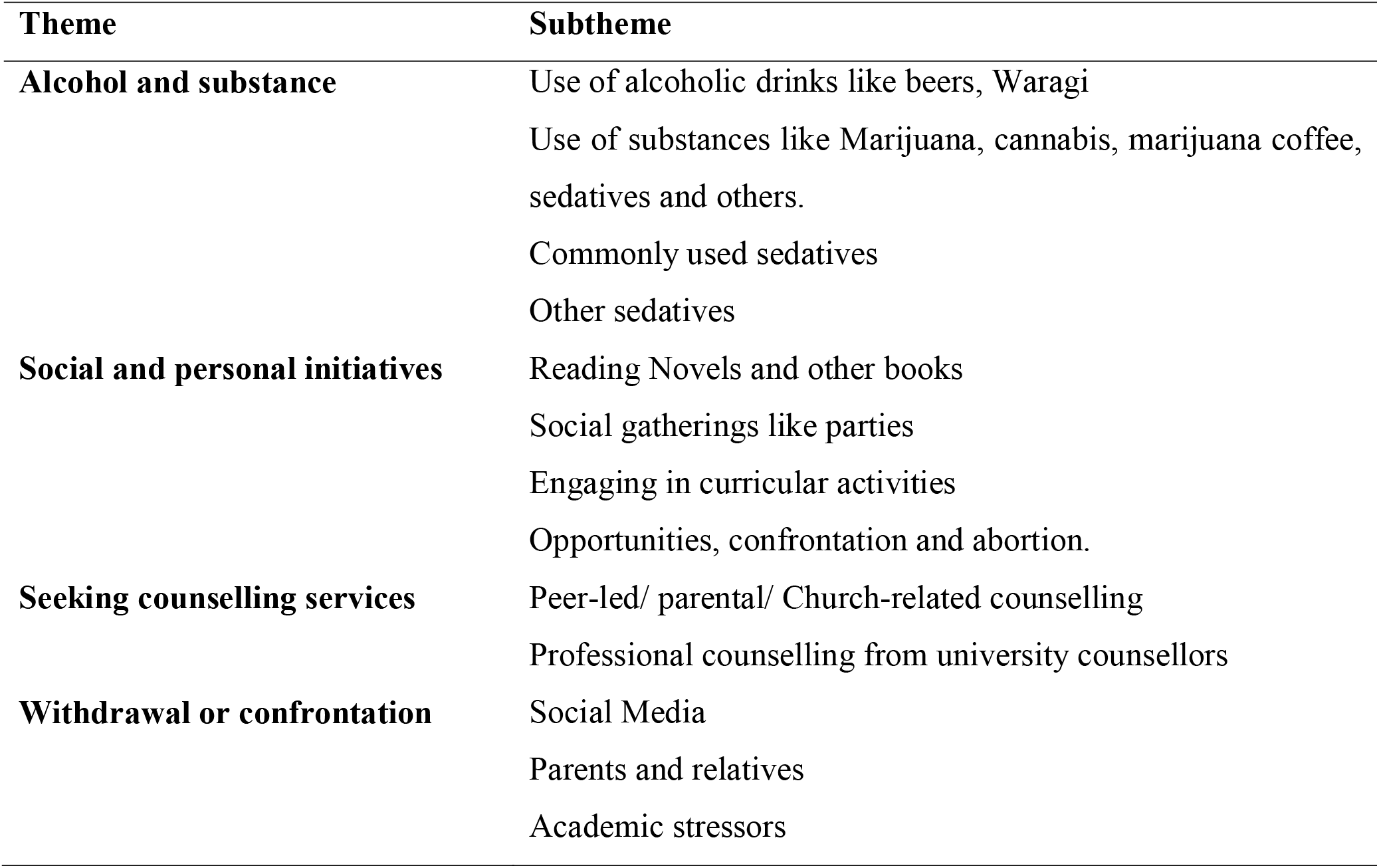
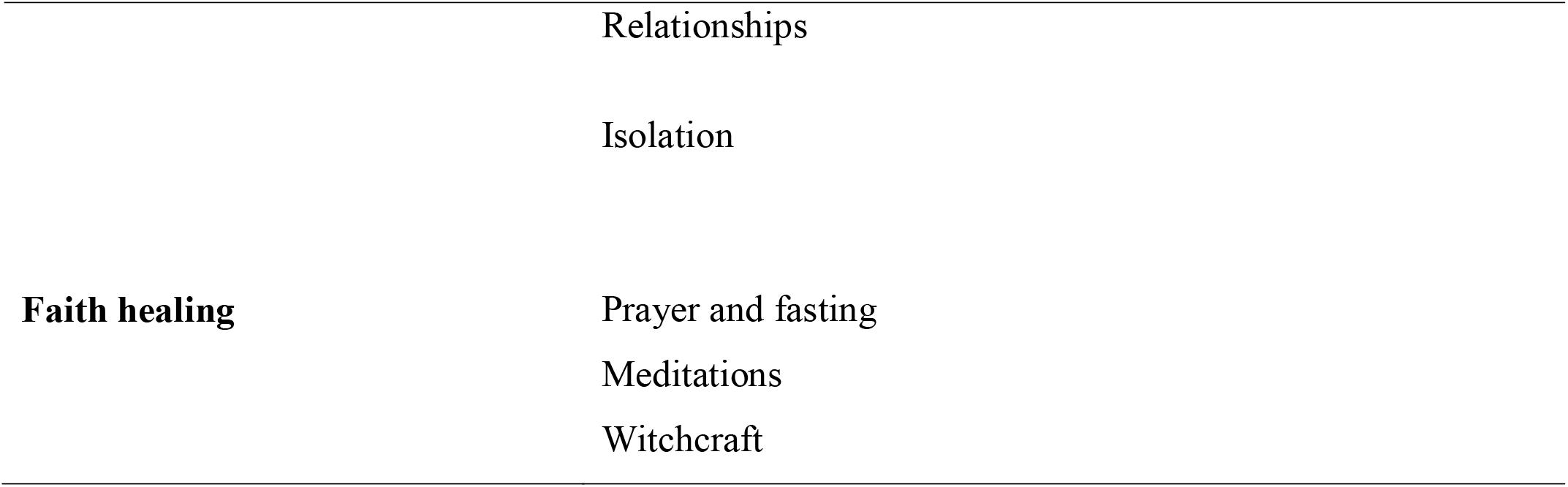
Thematic Areas of coping mechanisms among university students.

| Theme | Subtheme |
| --- | --- |
| **Alcohol and substance** | Use of alcoholic drinks like beers, Waragi |
|  | Use of substances like Marijuana, cannabis, marijuana coffee, sedatives and others. |
|  | Commonly used sedatives |
|  | Other sedatives |
| **Social and personal initiatives** | Reading Novels and other books |
|  | Social gatherings like parties |
|  | Engaging in curricular activities |
|  | Opportunities, confrontation and abortion. |
| **Seeking counselling services** | Peer-led/ parental/ Church-related counselling |
|  | Professional counselling from university counsellors |
| **Withdrawal or confrontation** | Social Media |
|  | Parents and relatives |
|  | Academic stressors |
|  | Relationships |
|  | Isolation |
| **Faith healing** | Prayer and fasting |
|  | Meditations |
|  | Witchcraft |

### Theme 1: Alcohol and substance use among students as a coping mechanism

The study identified use of alcohol and other substance as one of the coping strategies for undergraduate students at Busitema University Mbale and Tororo Compass. Most if the students acknowledged having taken a beer to solve their problems while some were victims of other drugs and substances like marijuana, and cannabis to mention but a few. Others preferred to use cheaper options like taking “coke” (a local soft drink) all the time yet a few of them afforded to take coffee often. Also some students reported using sedatives mainly for sleep related challenges related to mental disorders.

### Subtheme 1: Use of Alcoholic beverages/ drinks

Students revealed that they took some beers during the weekends and others on a daily basis after a hectic day. Some of the students at Mbale Compass informed us that the intense schedules and academic pressures prompted them to resort to taking some wines, beers and other alcoholic drinks to get a better feel of the day and gain energy to take on the mantle for the next day.

> *“…*..*I take beer, especially after a stressful day, we meet at some place in shawole with some colleagues and have some kind of fun*. ***FOE, Male, Year 3, BCT, 22-30 years)***
>
> *“I take wines, Smirnoff, but once in a while, Like at social events like parties, when I get a chance to go to clubs etc* ***(FHS, Female, Year 5, MBchB, 21-24 years)****”*.

### Subtheme 2: Substance use among students

The study revealed that some students admitted to taking illicit drugs and narcotics particularly cannabis and amphetamine when faced with mental health challenges. According to the students, it helped them “forgot their hardships”, “makes them feel high” (important) and facilitated their studies by improving their retentive memory.

> *“……some of us have been taking tramadol and others smoke weed and other stimulants because we think that when you smoke it, we shall forget your problems, you don’t think of that anymore* ***(FHS, Male, Year 3, MBchB, 21-24 years)****”*
>
> *“……Some of us smoke a lot and take alcohol for example I take a tusker just to keep myself calm and smoke some weed but once in a while with my friends…*.(***FOE, Male, Year 4***, ***WAR, 25-30 years)***.*”*
>
> ***“Yes***, *I used to use a lot of coffee and without it i thought I would not function, feel sad in class and tired all the time. If I fail to get coffee, I would get a Coca-Cola or Pepsi because I knew they had caffeine and it would make me energetic though it brought me problems because I took it in high amounts and took me back to the depression” crying… 2 minutes(****FHS, Female, Year 5, MBchB, 25-30years)***.*”*.

### Sub theme 3: Use of sedatives as a coping mechanism for students with mental health challenges

After a long working day, some students preferred to take sleeping pills and other forms of sedatives to calm them down and gain some sleep. The study identifies some types of sedatives used and when they were used by students.

### Subtheme 3 a: Use of sleeping pills

Some of the students we interacted with acknowledged the use of sleeping pills because of too much pressure, especially for those in the clinical years. Some preferred to take them to help them have a rest from the intense pressures at the university.

> *“I took an overdose of diazepam (I took a strip of diazepam) and the next day I found myself partially dead. Some people broke into my room and rushed me to the hospital and gave me an antidote and other treatment. I have been using it for a while to catch my sleep after trying other options and they disappointed me, (****FHS, Male, Year 3, MBchB, 21-24 years)*.*”***
>
> *“……One time I was forced to take sleeping pills that’s when I was rotating in internal medicine and ob-gyn, the pressure was too much and I had to look for an alternative*, ***(FHS, Female, Year 5, MBchB, 25-30 years)”***.
>
> *“I used to take sleeping peels “Diphenhydramine” all the time I wanted to have constructive sleep but woke up the following day when my friends were coming back from class* ***FHS, Female, Year 5, MBchB, 21-25 years)”***.,

### Subtheme 3 b: Other sedatives

The study also revealed that some students were using other sedatives like coffee, and taking a lot of coke to help boost their activity or slow it and also reduce or slow attention span.

> *“I would also take amitriptyline, I know it was a bad coping mechanism but it was one way to go*, ***(FHS, Female, Year 4, BNS, 25-30 years)***
>
> *“There is a ka drug I bought but I overslept and missed lectures that day and vowed not to take it again because I thought that when I sleep for long the anxiety will go but it didn’t, I felt better for some time and went back. I got it over the counter from one of the pharmacies around town*. ***(FOE, Male, Year 2, AMI, 21-25 years)***

### Theme 2: Social and personal initiatives

The study also revealed that students created their own personal initiatives to keep themselves mentally stable. The initiatives included reading novels, attending social events like parties, and going to clubs, while some opted to engage in curricular activities.

### Subtheme 1: Reading Novels and other books

To those who preferred reading novels, they did it to reduce on the anxiety and depression that they were experiencing at that time. Some of them informed us that whenever they felt suicidal impulses, reading the bible was the best alternative.

> *“I do read novels most times when I feel life has lost meaning and I encourage other students to do the same instead of crying whenever they are depressed*.***(FOE, Female, Year 3***,***BEE, 25-30 years)”***
>
> *“There are times when I had financial and family issues, I felt like killing myself but I resorted to reading some good chapters in the bible and realized that Jesus loves me, killing my self was not an option***(*FHS, Male, Year 2, MBchB, 21-25 years)***.

### Subtheme 2: Attending social gatherings like parties

Attending social gatherings like parties, going to night clubs as well as movie nights were also coping mechanisms identified among students at both campuses. Most of them informed us that when they went to club, they would engage in dancing the whole night and relieve the stress and pressures. Parties like medical dinners, cultural galas, introductions and weddings of their colleagues were also coping mechanisms as they helped to keep them away from the university pressure.

> *“Once in a while, I go to club el-tanjia when there are artists, I dance and kill the stress off as I wait for the next week and its problems* ***(FHS, Female, Year 1, BNA, 21-25 years)”***.
>
> *“I attend parties for our chapel and colleagues who are getting married which keeps me off my final year project for some time* ***(FOE, Male, Year4, BCT, 25-30 years)”***
>
> “We have a cultural gala in Mbale which I participate in and during that whole period, I forget about the stress of books, exams etc. ***(FHS, Male, Year 3, BNS, 21-25years)”***

### Subtheme 3: Engaging in curricular activities

The study found out that some of the students also resorted to engaging in co-curricular activities especially those at Tororo compass. Unlike students at Mbale compass whose engagement in co-curricular activities was limited because of lack of space, those at Tororo compass embraced it to solve their mental health challenges.

> *“Yes, I do football with friends every evening and this has helped me to avoid being alone and over thinking* ***(FOE, Male, Year DGE, 21-25years)”***.
>
> *“Yeah at our compass, we don’t have where to play football from however we have had some time to play at Mbale College of health sciences, this has made us get relieved a little* ***(FHS, Male, Year 4, BNA, 25-30 years)”***.

### Subtheme 5: Opportunities, confrontation and abortion

The confrontation was one of the mechanisms that students felt would help them solve their problems, especially misunderstandings that existed between them and their lecturers,

> *“For the Lecturer who segregates us in class, my dear…*.*ehhhh…*.*I went ahead and confronted that lecturer who felt like his tribe mates were more important than some of us, I know I did a wrong thing but the guy styled* ***(FHS, Male, Year 5, MBchB, 21-24 years)”***.

Some students believed in the opportunities ahead of them and utilized them optimally especially extensors who had lost their jobs yet they served as the only source of income for them and their families,

> *“Iam looking for an alternative job in Mbale and I got one in Abacus but it was in Kampala and yet it was hard for me to balance books and school. When I got another one, the opportunity was in Mbale and Arua and they would not allow me to choose the place mate site and they ended up posting me in Arua and I missed out just like that* **(*FHS, Male, Year 2, BNA, 25-30 Years)”***.

Regarding accommodation, some of them ended up renting around town although they felt unsecure. Since their hall of residence was not conducive, they paired up and decided to pay rent on a monthly basis, it was costly but far better than the hostels.

> *“We ended up going out as in renting but we did not have that good security like at bellodian and the rent was too much for us. I have friends who were robbed because they are renting and even with in bellodian that kept traumatizing them* ***(FHS, Female, Year 5, MBchB, 21-25years)”***.

Regarding some of the financial challenges that were identified among the students, parental support was sought and others sought scholarships from respective organizations. Some students carried out fundraising campaigns for their students who were struggling financially while the extensors continued looking out for job opportunities to support them financially.

> *“My fellow colleagues ran a fundraising campaign for me and we were in position to raise 850 K and for sure when they gave me that money, it was enough for me to heal psychologically, I felt like it was better than nothing* **(*FHS, Female, Year 1, BNS, 21-25 years)”***.
>
> *“I got a sponsor through a small organization called “blue house” in Kazo district and it took me up and we stayed there from P*.*5 up to s*.*6. Life was okay but it would take a toll on your emotional wellbeing* **(*FHS, Female, Year 2, MBchB 21-25 years)”***.
>
> *“I was forced to borrow money from colleagues and money lenders who have also become a headache to me* ***(FOE, Male, Year 2, MEB, 21-25 years)”***.

### Subtheme 6: Listening to music and socializing

The study also revealed that listening to music was one way of dealing with psychological distress among students. Some opted to make themselves busy as way of forgetting the challenges and hustles off the day.

> **“***I listen to Christian music all the time as long as I am not at school and it has helped me to deal with the suicidal behavior which I had (****FOE, Male, Year 1, AMI, 21-25years)”***.
>
> *“I am a very task-oriented person, so being in the ward busy would make me forget some of the things related to the problems I had*, ***(FHS, Female, Year 4, MBchB, 22-30 years)***

### Theme 3: Seeking counselling services

While as some students opted for the above coping strategies, counselling was also another strategy embraced mostly by students at both campuses despite the absence of enough counsellors. They used the alternatives like peer led counseling’s, parental counselling and others preferred to visit professional counsellors and psychiatrists or more help.

### Subtheme 1: Peer-led/ parental/ Church-related counselling

Seeking Peers, parents and church leaders-led counseling was identified as one of the coping strategies for some students. They felt comfortable sharing their problems or challenges with their fellow peers, church leaders and parents and this reflects to what an English man referred to as “a problem shared is a problem solved”

> **“***I just kept talking to my friends and trying the above alternatives for the medical issues but for counselling, it was for my personal issues*, ***(FHS, Female, Year 5, MBchB, 25-30 years)”***.
>
> *“I would also talk to my parents and they offered all the necessary support they got me some mentors and they kept with me all the time*, ***(FHS, Male, Year 3, BNA, 21-25 years)***
>
> *“I trusted my friends and parents especially when I felt like I was developing bipolar. I also got self-help from U-Tube channels*, ***(FHS, Female, Year 1, BNS, 21-25 years***,***)”***

### Subtheme 2: Professional counselling from university counsellors

Some of the students preferred to visit professional counsellors and psychiatrists for help. Some were just referred to the counsellors while others just felt like they need to meet them for a psychosocial support.

> *“I also talked to some staff and the university counsellor who gave me some talk therapy*, ***(FHS, BNA, Year 3, 31-35 years)”***
>
> *“Yeah, I talked to some but not my personal problems, I would tell them about academic issues because even when we used to tell them, they used to tell us that it’s the nature of medical school and you have to persevere*, ***(FHS, Female, Year 4, BNS, 25-30 years)***

### Theme 4: Withdrawal as a coping strategy

Some of the students decided to withdraw which were contributing to their mental unwell being like relationships, social media, some nagging relatives and friends and resorted to concentrate on their academics and what their gals were.

### Subtheme 1: Withdraw from social media

The study revealed that some of the students decided to withdraw from social media which had caused some mental health challenges to them, since social media is a collection of information, some ended up watching and reading messages and information that left them depressed and in so doing withdrawing from it was the only solution.

> *“I withdrew from all social media platforms and switched off my phones and we went for training and distanced myself from things that were taking a lot of my thoughts* ***(FHS, Female, Year 3, MBchB, 21-25 years)***.
>
> *“I turn off social media and withdraw which used to expose me to some funny messages and also changed companies as in the people I was associating with from people* ***(FOE, Male, Year 2, WAR, 21-25years)***.

### Subtheme 2: Withdrawal from parents and relatives

Some also opted to withdraw from some relatives and friends who had become problems to them and were a trigger to their mental unwell being.

> *“I distanced myself from people whom I thought had negative thoughts towards me* ***(FOE, Female, Year 4, WAR, 21-25 years)***.
>
> *“I avoid talking to people and that is why you can find me with heads on even while moving on the streets just to avoid talking to people*. ***(FHS, Female, Year 3, MBchB, 21-25 years)”***.

### Subtheme 3: Withdrawal from Academic Stressors

Withdrawal from leadership positions and other forms of academic stressors was also another coping strategy for some of the students at both campuses.

> *“Students ended up leaving the course and others while in year one decided to apply and join other universities because the administration was not supportive*,***(FOE, Male, Year 2, DCE, 21-25 years)”***.
>
> *“I had many leadership roles like class representative, choir leader, Rotaract treasurer, I was in women in the medical world etc but I withdrew from them and my life took another direction. At that time my BP normalized because it was unstable and I used to get palpitations* ***(FHS, Male, Year 5, MBchB, 25-30 years)”***

### Subtheme 4: Withdrawal from relationships

The decision to withdraw from relationships was a personal take by some of the students to relieve themselves from the relationships and their associated complications. Some found purpose in their academics and preferred to concentrate on their goal, ambitions and academic pursuits.

> *“For relationships, I decided to concentrate on the person I wanted to be and also withdrew from the relationships though you can see me pregnant but I won’t go for any other relationship, this was a lesson to me*. ***(FOE, Female, Yeah 4, WAR, 25-30 years)***.
>
> *“I withdrew from the relationship I had with some lecturer, went to church and prayed for the guy, my worry was a retake in his course unit but I read hard and passed****(FHS Female, Year 3, MBchB 21-25 years)”***.

### Subtheme 5: Isolation

Due to the stigma associated with mental illness, isolation, seclusion and avoidance of social contacts were the primary adaptations for some students at both compasses.

> *“When you let people know what you are going through, they will think you are mad. I just keep to myself lonely and avoid listening to people that’s why you see me with headsets all the time* ***(FHS, Female, Year 3, MBchB, 21-25 years)”***.

### Theme 5: Faith healing

The study identified another coping strategy which was seeking help from supernatural sources through prayer and fasting, meditation and witchcraft. It was surprising that some of the students at Tororo compass preferred to use witchcraft to revive their business in spite of the tight completion around them. Witchcraft was also identified as a solution to the restoration and maintenance of relationships by some of the students.

### Subtheme 1: Prayer and fasting

The study revealed that for most of the students amidst the hardships and struggles with psychological distress and other mental health challenges, prayer and fasting were the only strategies to help them cope. They believed in the biblical principles(16) of Philippians 4: 5-7 which says that **“**Do not be anxious about anything, but in everything by prayer and supplication with thanksgiving let your requests be made known to God and the peace of God, which surpasses all understanding, will guard your hearts and your minds in Christ Jesus” and Matthew 17:21 which says that “But this kind does not go out except by prayer and fasting.”

> *“Sometimes I would go to the chapel for prayers and praise and worship and also engage in some days of prayer and fasting,(* ***FHS, Female, Year 1, MBchB, 21-25 years)”***.
>
> *“I talked to my friends, especially those we missed, cried over it and gave up because we had it in mind that the lecturer just wanted to punish us, so I cried and cried and gave up*.***(FHS, Female, Year 5, MBchB, 21-25years”)***.
>
> *“I used to pray a lot and talk to God and it could make me better with both depression and anxiety*. ***(FOE, Male, DAG, 25-30 years)”***

### Subtheme 2: Meditation

Some of the students were actively involved in meditation which helped them to keep their mental health stable at all times.

> *“I went through guided meditation and learnt new habits of controlling myself and read books “No one can affect your moods unless you have accepted” so things became okay* ***(FOE, Male, Year 2, AMI, 21-25 years)***.
>
> *“I was going through a brainwash, I could get something and meditate like for 30 minutes and avoid thoughts that would be running in my mind* ***(FHS, Male, Year 2, BNA***, ***21-25 years)***..

### Subtheme 3: Witchcraft

The study also revealed that some of the students at one of the compasses were involved in witchcraft. It was surprising that some of the students at Tororo compass preferred to use witchcraft to revive their business in spite of the tight completion around them. Witchcraft was also identified as a solution to the restoration and maintenance of relationships by some of the students.

> *“When my business was not progressing well and it was my source of tuition, I was advised by colleagues to try witchcraft since most of the colleagues in business were doing so and it worked for me, I actually cleared all my tuition* ***(FOE, Male, Year 2, BEE, 21-25 years)***.
>
> *“I was forced to use witchcraft because I was told that it was the only remedy that would return my girlfriend. I had already done kukyala and waited for us to finish school and have a family but that guy snatched her and it gave me sleepless nights for almost a week*, ***(FOE, Male, Year 3, WAR 25-30 years”***.
>
> *“Here in shawole, most people use witchcraft in such scenarios, I also gave it a try though no results yet* ***(FOE, Male, Year 4 BCT, 25-30 years”***.

## Discussion

This study aimed at exploring the major copying strategies used by undergraduate students at Busitema University who were diagnosed with mental health disorders. Our findings indicated that students at both compasses had different copying strategies however there were some which were similar or common among all like faith healing, use of alcohol and substances, withdrawal among others. Copying mechanisms like use of alcohol and other substances was associated to the heavy workloads, the pressure for exams and final year projects and the poor and learning environments. The study found a lot of social, economic and personal related coping strategies among students at both campuses of Busitema University. While as others felt like substance and alcohol use would relieve them from the academic pressures and stress at school, other students used personally led initiatives like reading novels and listening to music. Our findings were not far from what a study conducted among students in in other countries like Nigeria and Ghana(15). Most students with mental illness were highly stigmatized and sometimes regarded as cursed, mad or possessed and thus often suffered in silence and isolation because mental illness has spiritual connotations, and those ‘afflicted’ sought divine intervention from all religious fonts like churches and mosques. In this study, seeking spiritual intervention was one of the coping strategies embraced by students at both campuses which was not different from a study conducted in turkey(9). Although witchcraft and curses are often believed to cause not only mental illness but general misfortunes in life, students at one of the campuses opted to use them to solve their financial and relationship problems which related to the general oracle of witchcraft(17). Religious support was known to influence hope, life satisfaction and emotional well-being which also appeared in a study conducted in Poland during the COVID-19 pandemic. (6). Although some of the alcoholics and substances were not acceptable for students, the use of stimulants and amphetamine such as tramadol and cannabis particularly marijuana to deal with stressors was common among students at both campuses and similar findings were discovered by Kamran Sattah in his scoping review(3). Our findings also revealed that receiving psychosocial support through peer-led, parental and trained counsellors from friends as a copying strategy helped to reduce the prevalence of mental disorders among students at both campuses and this was not very different from the finding of a similar study conducted among college students in Bangladesh (8). Having guidance and counselling services at both compasses is a laudable innovation that students would embrace however, we noted that the purpose of counselling was derailed by concerns related to less trust, confidentiality and the absence of enough counsellors, especially female counsellors both compasses. Since most of the students had a negative attitude towards counselling(18), creating awareness on guidance(12) and counselling and employing trained counsellors(19) could enhance access to counselling services by students. Mental health education should be introduced(20) to all students at all campuses of Busitema University and cascaded to other institutions as well.

### Strengths and limitations

The study was able to capture responses from all the students who were diagnosed with mental disorders at both compasses.

The only limitation was related to less time allocated for the study.

## Data Availability

The datasets used and analyzed during the current study are available from the corresponding author on reasonable request.

## Conclusion

Mental health challenges among students are a level higher in most institutions of higher learning however their coping strategies amidst such situations needed to be looked through and given much attention. Some are self-thought-through strategies while others are university-led interventions. The need to streamline such intervention is necessary for the betterment of the mental health status of students.

## Acknowledgement of the funding body

This study was fully supported by the Busitema University Research and innovation fund grant 3/DGSRI/22. We are grateful to the administration of Busitema University and student leaders of Busitema and Mbale campuses who supported us throughout the data collection process.

## Author contributions

EKK conceived the idea, wrote a proposal with JK and JLM. EKK and JK oversaw and participated data collection, analysis and interpretation of results, drafted manuscript and proofread integrated all co-authors contributions. JLM, AGN, FO, EW, COA, AN, MLRM, RK,AO,KN and PW supported with proposal writing, data collection, analysis and interpretation of results and proof read all versions of manuscript and provided additional technical support to the team. All authors have read and approved this manuscript prior to submission.

